# Did Norwegian adolescents suffer more violence and sexual abuse during the Covid-19 pandemic? Violence and sexual abuse rates the year before the pandemic compared to rates one year into the pandemic

**DOI:** 10.1101/2022.04.26.22274316

**Authors:** Else-Marie Augusti, Mia Cathrine Myhre, Tore Wentzel-Larsen, Gertrud Sofie Hafstad

## Abstract

**Background:** The Covid-19 pandemic is a public health crisis which may cause unintended additional societal costs such as child maltreatment. Considerable concern is raised as to whether the pandemic has led to an increase in violence and sexual abuse against children.

**Objective:** The present study objective is to provide rates of violence and sexual abuse against adolescents the year before the pandemic compared to one year into the pandemic.

**Participants and setting:** Two samples of Norwegian 12-16-year-olds were approached. A representative pre-pandemic sample of 9240 adolescents (*M* age= 14.11), and a sample recruited one year into the pandemic resulting in 3540 responses (*M* age *(SD)* = 14.5).

**Methods:** An online survey was administered during school hours including established measures of violence and sexual abuse exposure. Sociodemographic characteristics were assessed.

**Results:** There was 1.4 percentage point increase in sexual abuse by an adult, and a 3.9 percentage point decrease in psychological violence by a parent during the pandemic compared to the year before the pandemic. Otherwise, violence and sexual abuse rates remained stable across these two time periods. Risk factors for violence and sexual abuse were amplified during the pandemic.

**Conclusion:** Norway, a high-income welfare state, imposed measures to counteract the burden of the pandemic mitigation actions for adolescents. This might partly explain the absence of the feared increase in violence towards adolescents. The disproportionate risk for violence and sexual abuse for some groups of adolescents is however concerning, and should be followed up over time.

---

The measures adopted to mitigate one public-health crisis can potentially cause other unintended negative effects on public health. This is the case for the Covid-19 pandemic and mitigation measures implemented, causing what some consider the perfect storm for domestic violence and child abuse (Usher et al., 2021). Stress and toll on families during this period have been highlighted across the world. Parents’ efforts to combine work-related stressors, financial hardship, worries for the pandemic, and the higher burden of parenting imposed by the enforcement of home-schooling and home daycare can tax family life (Griffith, 2020). As such, concerns have also been raised on an increase in rates of violence and abuse towards children. However, few studies have established rates of violence and abuse under the pandemic, and even fewer use a design allowing to assess stability or change in rates of violence against children from before to during the pandemic. A few exceptions exist, for instance studies based on historical data from children’s emergency departments (ED) published since the onset of the pandemic. These studies have in general shown a decrease in injury-related ED visits due to violence (De Boer et al., 2021; Sokoloff et al., 2021), but some also indicate an increase in the more severe types of violence against children (De Boer et al., 2021; Morris, Rogers, Kissmer, Du Preez, & Dufourq, 2020). Studies from administrative records from the child welfare services reported drops in reports during the first phases of the pandemic, Spring and Summer 2020. Rates of reports bounced back to pre-pandemic levels during the later months of 2020 (Barboza, Schiamberg, & Pachl, 2020; Baron, Goldstein, & Wallace, 2020; Bhopal, Bagaria, Olabi, & Bhopal, 2021; Katz et al., 2021; Rapoport, Reisert, Schoeman, & Adesman, 2021). In all studies, the decreases identified were not taken as a signal of an actual drop in incidence, but rather a testament of the cruical role schools and health care services represent in the detection of child maltreatment (Thomas, Anurudran, Robb, & Burke, 2020). However, as revealed by a recent systematic review (Marmor, Cohen, & Katz, 2021), self-report data on child abuse during the pandemic is lacking it is therefore paramount to surveil self-reported child abuse numbers to accompany information from official records (Marmor et al., 2021).

Furthermore, the present pandemic has shed light on groups living in disproportionate risk for negative effects of the mitigation measures imposed. Previous research has shown that increasing the burden of parenting in households with existing risk factors increase the propensity for negative outcomes not only for the parents, but also for the children in these families (Griffith, 2020). Underscoring this, studies from the current pandemic has shown that in families with a history of harsh parenting and violence and/or mental distress, the risk for abuse during the pandemic increased (Brown, Doom, Lechuga-Peña, Watamura, & Koppels, 2020; Lawson, Piel, & Simon, 2020; Wolf, Freisthler, & Chadwick, 2021). Based on a different subsample from the present study, we found increased odds for experiencing violence and abuse during the pandemic for previously identified at risk populations such as children residing in families with financial difficulties, where parents suffer from mental illness, drug abuse or in single-parent households (Augusti, Sætren, & Hafstad, 2021). Self-reported life-time exposure to violence or abuse one year prior to the pandemic, was the most prominent risk factor for abuse experiences during the pandemic in this sample (Augusti et al., 2021).

## The present study

Norway has been affected by the pandemic by intrusive measures in the attempt to significantly slow down the spread of the virus. As of fall 2021, the mitigation measures implemented seem to have had the wanted effect, minimizing deaths and severe illness in the population. However, the mitigating measures have had their toll on children and families, especially in the greater metropolitan area of the capital city of Oslo. For longer periods of time during Fall 2020 through April 2021, Oslo and surrounding areas have effectuated remote-teaching and home-schooling for adolescents in middle- and high schools. Restaurants, cafeterias, and shops, except from grocery stores and pharmacies, were closed during the Winter of 2021. Indoor sports and leisure activities were terminated for a significant amount of time for the oldest age groups (12-18-year-olds). In addition, quarantine and isolation were imposed if family members were exposed to or infected with the virus. Not until late spring 2021 did schools, sport and leisure activities, shops and restaurants reopen in the capital city of Norway and surrounding areas. For most other parts of the country, the situation was strikingly different, imposing few to none restrictions and reporting significantly lower rates of illness and deaths than that reported in the Oslo region. Nevertheless, other parts of Norway also experienced lock-downs during times of locally high incidence of Covid-19-infections. Moreover, several measures resulted in restricted access to after-school activities and reduced school hours. Additionally, unemployment rates were high across the country directly affecting the adult population, and indirectly affecting children and families.

The present study builds on a longitudinal data set with pre-pandemic and pandemic rates of child abuse in a large, representative sample of Norwegian adolescents between 12-16 years of age. This enables comparison of abuse rates the first year of the pandemic to the year before the pandemic in Norway. In addition, risk factors associated with exposure to child maltreatment will be assessed at both measurement points in order to address the previously documented disproportionate risk for violence and abuse exposure in some groups of children.

## Methods

### Participants

The participants represented middle school students throughout Norway. They were all students attending mainly public schools, although a few private schools were also included. Two samples, partly overlapping, are included in the present study. The samples were drawn from the same middle schools, including participants aged 12-16 years. First data collection (January 2019) about one year before the COVID-19 pandemic comprises a sample of 9240 adolescents (*M* age *(SD)* = 14.11 (.88) years, 4592 47 % girls) representative for the Norwegian youth population (Hafstad et al., 2020). The last data collection (June 2021) one year into the COVID-19 pandemic generated 3540 responses (*M* age *(SD)* = 14.5 (.96) years, 1675 47 % girls). The majority of participants were born to parents from Norway or other Nordic countries. Most participants reported that they were living with both parents and that their parents were employed; a minority of adolescents reported perceived family affluence concerns. Additionally, when measuring family risk factors, a minority of the participating students had previously lived or were currently living with caregivers who had mental health problems, problems with alcohol or drug abuse, or who had been or were currently incarcerated. For details on the included study samples, please see Table 1.

**Table 1.** Demographic characteristics of the two samples included in the present study.

|  |  | Pre-pandemic<br>(2019) |  | Pandemic<br>(2021) |  |
| --- | --- | --- | --- | --- | --- |
|  |  | % | n | % | n |
| Gender | Girls | 47.1 | 4592 | 47.3 | 1675 |
|  | Boys | 46.5 | 4538 | 50.3 | 1781 |
|  | Gender non-confirming | 0.6 | 63 | 1.8 | 63 |
| Living arrangements | Living with both parents | 70.3 | 6458 | 69.3 | 2429 |
|  | Single-parent household | 29.7 | 2725 | 30.7 | 1077 |
| Family functioning | Living with parents without mental health or drug abuse problems | 76.6 | 7478 | 80.4 | 2846 |
|  | Unsure about parental mental health problems or drug abuse | 4.6 | 164 | 10.6 | 377 |
|  | Living with parents with mental health or drug abuse problems | 6.6 | 644 | 6.6 | 234 |
| Ethnicity | Norwegian or Nordic family origin | 68.7 | 6707 | 72.1 | 2554 |
|  | European family origin | 8.2 | 799 | 9.0 | 317 |
|  | Non-European family origin | 16.7 | 1532 | 16.3 | 576 |
| Perceived family affluence | Non-poor perceived family affluence | 89.9 | 8773 | 94.3 | 3337 |
|  | Poor perceived family affluence | 3.6 | 351 | 3.7 | 131 |
| Geographical location | Living in Oslo metropolitan area during the pandemic | - | - | 43.8 | 1541 |
|  | Living outside the Oslo metropolitan area during the pandemic | - | - | 52.0 | 1830 |

**Table 2.** One-year prevalence rates of violence and abuse before and during the pandemic.

|  | Pre-pandemic rates |  | Pandemic rates |  | Diff. | CI* |
| --- | --- | --- | --- | --- | --- | --- |
|  | % | n | % | n |  |  |
| Psychological abuse | 12.0 | 1077 | 8 | 251 | -3.9 | -5.1, -2.8 |
| Physical abuse | 7.1 | 638 | 5.9 | 201 | -1.2 | -1.8, 0.04 |
| Sexual abuse by an adult | 3.1 | 284 | 4.5 | 154 | 1.4 | 0.6, 2.1 |
| Sexual abuse by a peer | 15.0 | 1341 | 13.9 | 473 | -1.1 | -2.4, 0.3 |
| Witnessing domestic violence towards mother | 8.6 | 760 | 7.6 | 254 | -1.0 | -2.0, 0.1 |
| Witnessing domestic violence towards father | 7.3 | 640 | 5.9 | 194 | -1.4 | -2.4, -0.5 |
| At least one abuse experience during the last year | 27.8 | 2713 | 25.7 | 909 | -2.12 | -3.8, -.04 |

Note. \* = CI for the difference in percentage points between pre-pandemic and pandemic prevalence.

### Procedure

A total of 70 schools were included in the UEVO-study (pre-pandemic;(Hafstad, Sætren, Myhre, Bergerud-Wichstrøm, & Augusti, 2020). These schools were re-approached and asked to administer the web-based survey to their student body in 8th through 10th grade one year into the pandemic. Due to the extraordinary situation facing schools and society in general during the COVID-19 pandemic outbreak, only approximately 50 % (n = 35) of the approached schools agreed to take part in the last data collection.

For both time points, participating schools administered the survey during school hours. The web-based survey took approximately 40 minutes to complete. A short animated video about the study and information about the ethical principles of voluntary participation, confidentiality, and the right to withdraw at any time during the study without having to give a reason preceded each data collection.

After data collection, the two data sets were merged. Merging was done based on participant personal-ID number retrieved automatically when adolescents consented to participate in the study. The study was approved by the Regional committee for ethics in medical and health research in the Southeastern region of Norway (Case #2018/522). All participants provided informed digitally written consent, no parental consent was required for the adolescents to participate. For a detailed description of the study procedure and ethical considerations please see (Hafstad et al., 2020).

### Measures

#### Background questions

Participants reported their age in years and gender as either boy, girl or non-binary. Non-binary respondents were excluded in separate analyses for gender, due to the small sample size of this group. To assess parents’ country of origin participants were asked to indicate whether their mother and father, respectively, were born in Norway or a Nordic country (0), a European country (1) or a country outside Europe (2). Responses to these questions were combined into a composite variable on parents’ country of origin (both parents born either in Norway or the Nordic countries (0), at least one parent born in a European country other than a Nordic country (1), or at least one parent born in a country outside of Europe (2)).

Perceived family affluence was reported as 1) whether the adolescent experienced the family as having sufficient economic means to buy necessary goods; and 2) whether the adolescent had to decline after-school activities due to family finances. The first question was rated on a 4-point scale from 0 (completely agree) to (3) completely disagree, and the latter was rated on a four-point scale from 0 (never) to 3 (often). A perceived family affluence dichotomized composite score was derived based on responses to these two questions; if 2 or 3 was indicated on either of the two questions, a “low perceived family affluence score” (1) was allocated to that individual. Lastly, when measuring violence and abuse rates during the pandemic, we assessed whether the adolescent lived in close proximity to the metropolitan area of Oslo (plus or minus one hour drive). This served as a proxy to assess differences in rates depending on the burden of illness and mitigating measures imposed, as the Oslo metropolitan area faced more illness and prolonged periods of restrictive measures.

#### Family risk factors

Parents’ problems related to mental health, alcohol and drug misuse and incarceration were measured on a three-point scale (0 – No, 1 – Yes, 2 – Unsure).

#### Last-year violence and abuse prevalence

Based on a set of questions regarding abuse experiences, a follow-up question as to whether any of these experiences had happened during the last year, past 12 months, was used for the present analyses. The question regarding whether this had happened during the last year was posed after each set of questions described below, used to assess different types of abuse, and only if the participant yielded at least one experience of physical abuse by a caretaker, witnessing domestic violence at home, or sexual abuse by a peer or adult. More than one incidence of psychological abuse had to be endorsed in order to be counted as psychological abuse during the past year.

#### Physical and psychological abuse

Questions about physical and psychological abuse were inspired by the Parent-Child Conflict Tactics Scale (PCCTS; Straus, Hamby, Finkelhor, Moore, & Runyan, 1998) and adjusted to the present study population in line with other Nordic surveys on child abuse and neglect (Jernbro & Janson, 2017; Jernbro, Svensson, Tindberg, & Janson, 2012). Six questions of physical abuse spanned experiences such as pinching, pulling hair, spanking, beaten with a fist, an object, beaten up or being kicked. In addition, a last question was included to measure whether they had been exposed to other forms of physical violence. All questions were answered on a 4-point rating scale from never (0) to several times (3). Cronbach’s alpha for the full scale was .78. Psychological abuse was measured using six questions also inspired by the PCCTS (Straus et al., 1998). Questions about being ridiculed, threatened, locked in a room, or locked out of the house were all categorized as psychological abuse, yielding an alpha of .75. All questions were contextualized to capture abuse experiences imposed by caretakers. For psychological abuse, more than one incidence had to be endorsed in order to be considered as such.

#### Witnessing domestic violence

Witnessing domestic violence directed towards the mother, father, or a sibling respectively, was also administered. These questions were the same as those used in the Norwegian survey on child maltreatment among 18-year-old youth attending Norwegian high schools (Hafstad et al., 2020). Items covered witnessing psychological or physical violence against the child’s parent(s). Additionally, the children could indicate whether they had witnessed other types of violence against their parents. Items were rated on a 4-point scale from 0 (never) to 3 (several times). Witnessing violence against a sibling was assessed with the same 4-point scale.

#### Sexual abuse

Experiences of sexual abuse were assessed using four questions describing sexual transgressions involving being exposed to someone’s private parts, being touched on one’s own private parts or touching another person’s private parts, having intercourse with someone against one’s will, or other acts of a sexual nature. The same questions were posed twice, once pertaining to adults and once to peers, respectively. All questions were rated on a 4-point rating scale ranging from never (0) to several times (3), with a Cronbach’s alpha of .87.

### Statistical Analyses

Descriptive statistics are presented as percentages, means (*M*), and standard deviations (*SD)*. Prevalence rates of violence and abuse the year before the pandemic and the year of the pandemic were compared. To assess differences between pandemic and pre-pandemic times, bootstrap bias corrected and accelerated (BCa) confidence intervals for the differences were computed based on 10 000 bootstrap replications. We conducted logistic regression analyses to assess associations between known risk factors for abuse from caregivers (e.g., single-parent household, low socioeconomic status, immigrant background, parents’ mental health problems, alcohol and drug misuse, child’s gender) one year before the pandemic and during the first year of the pandemic. Physical, psychological and sexual abuse, and witnessing domestic violence were entered as dichotomized outcome variables in the logistic regression analyses. All analyses were conducted in SPSS version 26 (IBM Corp, released 2019), except the bootstrap procedure which was run in the R (1.4.116) package boot.

## Results

### One-year prevalence of violence and abuse before compared to during the Covid-19 pandemic

In all, rates of self-reported exposure to psychological and physical violence remained stable or declined somewhat, when measured the year before the pandemic compared to the first year of the pandemic. Sexual abuse perpetrated by an adult increased by 1.5 percentage points in 2020 compared to the year prior to the pandemic (2019), while we did not see evidence for an increase in peer sexual abuse. The results also indicate a reduction in psychological violence the first year of the pandemic, compared to the year before the pandemic outbreak. However, in all the differences detected were small, and confidence intervals (CI) for the differences indicate no significant change between the two time points assessed.

### Demographic differences in exposure

Girls reported a higher prevalence of violence and abuse exposure during the pandemic compared to boys. Interestingly, boys had a significant reduction in all types of violence during the year of the pandemic compared to what was reported the year prior to the pandemic. Whereas the rates for girls were stable across the two years of measurement (Table 3).

**Table 3.** One-year prevalence of violence and abuse before and during the pandemic for boys and girls respectively.

|  | Boys |  |  |  |  |  | Girls |  |  |  |  |  |
| --- | --- | --- | --- | --- | --- | --- | --- | --- | --- | --- | --- | --- |
|  | Pre- |  | Pandemic |  | Diff | CI | Pre- |  | Pandemic |  | Diff | CI |
|  | pandemic |  |  |  |  |  | pandemic |  |  |  |  |  |
|  | % | n | % | n |  | 95% | % | n | % | n |  | 95 % |
| Psychological abuse | 9.3 | 413 | 4.9 | 84 | -4.4 | -5.8, -5.1 | 14.4 | 655 | 11 | 179 | -3.4 | -5.3, -1.6 |
| Physical abuse | 6.6 | 294 | 3.8 | 66 | -2.8 | -3.9, -1.6 | 7.3 | 330 | 7.6 | 125 | 0.3 | -1.1, 1.8 |
| Sexual abuse by an adult | 2.1 | 94 | 2.2 | 38 | 0.1 | -0.7, -0.9 | 4.0 | 181 | 6.8 | 111 | 2.7 | 1.4, 4.1 |
| Sexual abuse by a peer | 9.7 | 426 | 5.5 | 94 | -4.2 | -5.6, -2.8 | 20 | 898 | 22.4 | 364 | 2.4 | 0.1, 4.7 |
| Witnessing mother being exposed DV | 7.1 | 309 | 5.5 | 93 | -1.6 | -2.9, -0.3 | 9.9 | 440 | 9.7 | 154 | -0.2 | -1.9, 1.4 |
| Witnessing father being exposed DV | 6.7 | 290 | 5.1 | 85 | -1.6 | -2.9, -0.3 | 7.9 | 343 | 6.6 | 103 | -1.3 | -2.7, 0.2 |

**Note.** \*DV: Domestic violence.

Violence and abuse rates did not differ significantly depending on whether the adolescents were living in a densely populated metropolitan area, with more strict and enduring lock-down measures, during the pandemic as opposed to living in areas that are more rural or less populated cities (Table 4). In fact, peer sexual abuse was reported significantly less in the Oslo metropolitan area compared to the rest of the country. This finding indicates that more social distancing might have acted as a protective factor for peer sexual transgressions.

**Table 4.** The Prevalence of Different Types of Violence and Abuse in the Metropolitan Area of Oslo Compared to the Rest of the Country during the first year of the COVID-19 pandemic.

|  | Oslo Metropolitan Area |  | Norway outside the metropolitan area |  | Diff. | CI |
| --- | --- | --- | --- | --- | --- | --- |
|  | % | n | % | n |  |  |
| Psychological abuse | 8.6 | 129 | 7.3 | 129 | 1.3 | -1.5, 1.7 |
| Physical abuse | 5.7 | 86 | 5.6 | 100 | 0.1 | -1.4, 1.7 |
| Sexual abuse by an adult | 4.5 | 67 | 4.4 | 78 | 0.1 | -1.5, 1.7 |
| Sexual abuse by a peer | 13.5 | 201 | 14.5 | 255 | -0,9 | -1.5, 1.7 |
| Witnessing mother exposed to DV* | 7.6 | 111 | 7.7 | 133 | -0.1 | -1.5, 1.7 |
| Witnessing father exposed to DV | 5.9 | 86 | 5.7 | 97 | 0.3 | -1.4, 1.7 |

**Note.** \*DV: Domestic violence

### Risk factors associated with violence and abuse before and during the Covid-19 pandemic

Logistic regressions showed that the same risk factors emerged as significantly associated with violence both the year before and the first year of the Covid-19 pandemic. Low perceived family affluence and reports that parents had current or previous problems related to mental health, alcohol and drug misuse and incarceration presented with significantly increased odds for violence and abuse experiences at both time points (see Tables 5 and 6). There was no indication of some risk factors being more potent than others when predicting violence and abuse exposure, during as compared to before the pandemic. Although the odds for experiencing violence and abuse for most risk factors were somewhat higher during the first pandemic year, compared to the year prior to the pandemic onset, the confidence intervals were still overlapping and thus not clearly representing a significant difference in risk.

**Table 5.** Logistic Regressions on the Association between Risk Factors for Violence and Abuse the Year before the COVID-19 Outbreak

|  | Psychological<br>abuse | Physical abuse | Witnessing DV<br>towards mother | Witnessing DV<br>towards father | Sexual abuse by<br>an adult | Sexual abuse by a<br>peer |
| --- | --- | --- | --- | --- | --- | --- |
|  | OR (CI) | OR (CI) | OR (CI) | OR (CI) | OR (CI) | OR (CI) |
| Gender, boy | 0.61 (0.52,<br>0.71)** | 0.98 (.83, 1.16)** | 0.72 (.62,.85)** | 0.90 (.76,1.06) | 0.55 (.43,.71)** | 0.45 (.39,.51) |
| Perceived family affluence | 2.10 (1.57,2.81)** | 2.21 (1.62,3.01)** | 1.46 (1.03,2.06)<sup>†</sup> | 1.46 (1.00,2.12) | 1.72 (1.08,2.72)* | 1.54 (1.17,2.04) |
| Parents' country of origin<br>– Europe (non-Nordic) | 1.05 (.81,1.36) | 1.23 (.93,1.62) | 0.82 (.62,1.09) | .77 (.56,1.04) | 1.03 (.67,1.57) | .72 (.57,.90)<sup>†</sup> |
| Parents' country of origin<br>– outside of Europe | 1.16 (.95,1.41) | 1.12 (.90,1.39) | .79 (0.64-,0.99)<sup>†</sup> | .61 (.48,.49)** | .77 (.54,1.10) | 0.60 (.50,.72) |
| Single-parent household | 1.02 (.87,1.20) | 1.09 (.91,1.31) | 0.66 (.55,.79)** | .55 (.45,.67)** | 1.12 (.86,1.47) | 1.24 (1.09,1.42) |
| Family functioning | 2.64 (2.25,3.09)** | 3.07 (2.55,3.69)** | 2.97 (2.50,3.54)** | 3.32 (2.75,4.00)** | 3.29 (2.52,4.31)** | 2.15 (1.86,2.49)** |

Note. <sup>†</sup> p>.05; \* = p>.01; \*\* = p>.001. DV: Domestic violence; Family functioning: Parents' mental health problems, parents' substance abuse

**Table 6.**
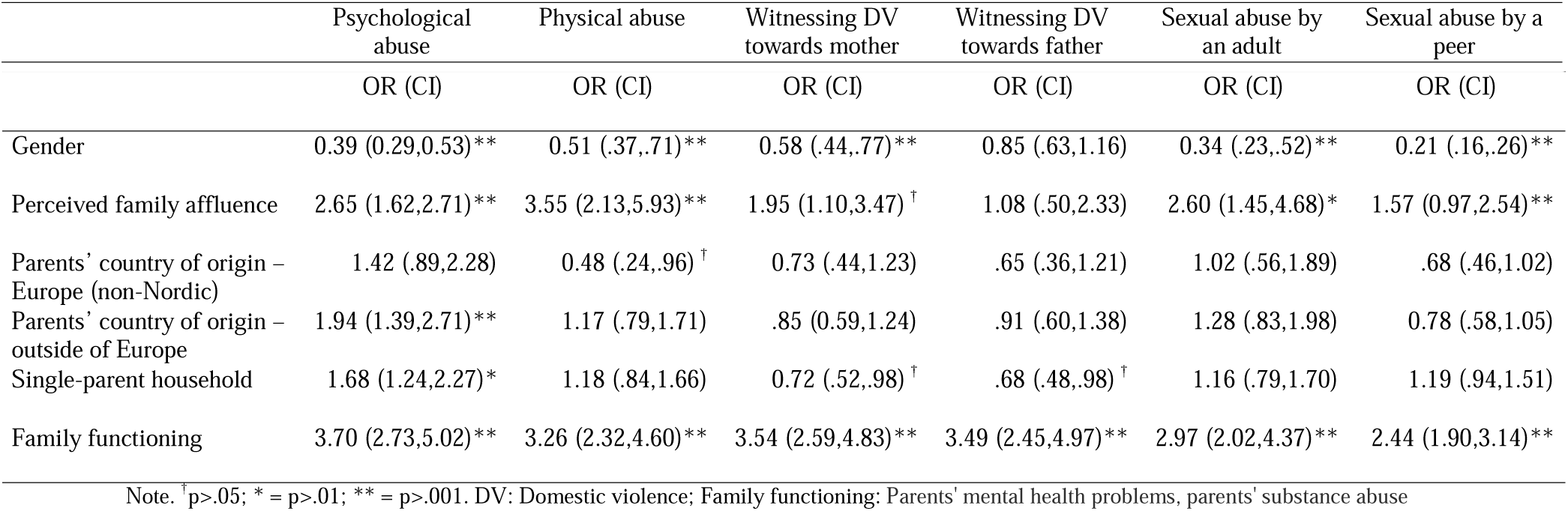
Logistic Regressions on the Association between Risk Factors for Violence and Abuse during the first year of the COVID-19 Outbreak

## Discussion

As one of very few studies to date, we have been able to compare last-year prevalence rates of self-reported physical violence, psychological violence and sexual abuse one year before to one year into the Covid-19 pandemic in a representative sample of adolescents. We found a small significant decrease in psychological abuse and a marginal, but significant increase in sexual abuse perpetrated by an adult. Apart from that, rates of adolescents’ self-reported violence perpetrated by a parent or sexual abuse perpetrated by a peer were stable across this time period. About one quarter of the adolescents participating in the study reported having experienced at least one incidence of the types of abuse mapped in the present study at both time points. Most frequently reported was sexual abuse by a peer, followed by psychological abuse by a caregiver. Sociodemographic characteristics of the sample did partly nuance this overall picture. Boys reported significantly less exposure to all types of violence and abuse during the pandemic, compared to the year before, whereas girls’ exposure remained about unchanged across the two measurement points. Living in a densely populated metropolitan area with more restrictive and prolonged measures to slow down the spread of the virus was not significantly associated with an increase in violence and abuse rates. A slight increase in psychological violence at home was however reported by adolescents living in areas with more restrictions compared to those living in less densely populated areas with fewer restrictions. A lower proportion of adolescents living in the metropolitan area of Oslo, reported peer sexual abuse compared to the rates reported by adolescents living outside the Oslo metropolitan area.

Partly in contrast to what has been raised as a concern in commentaries and papers published (Collin-Vézina, Brend, & Beeman, 2020; Griffith, 2020; Hoffman & Miller, 2020), rates of violence and abuse did not increase substantially in this representative sample of Norwegian adolescents. Moreover, the risk factors for being a victim of violence and/or abuse remained virtually the same during the pandemic as those identified the year before the pandemic. As numerous studies have described, child abuse is disproportionately affecting children in families with several other stressors present (Patwardhan, Hurley, Thompson, Mason, & Ringle, 2017). Scientists on child development and child welfare, and organizations working with children and families, were particularly concerned about children living under at risk conditions when the pandemic stroke (Marmor et al., 2021). In line with this concern, the relative risk for experiencing child abuse in some risk groups was somewhat higher during the pandemic compared to the year before the pandemic in the present study. However, the pattern of risk factors was close to identical before and during the pandemic. These were factors such as children’s perceived family affluence and parental psychosocial stressors such as mental and substance abuse related illness. Girls were also disproportionately affected, as shown both in the rates of violence and abuse reported especially during the pandemic, but also before the pandemic. However, the vulnerability associated with being a girl was more pronounced during the pandemic compared to the year before.

Sexual abuse perpetrated by an adult did, also, increase somewhat during the pandemic. The slight increase registered for child sexual abuse resonates well with findings from police authorities in Norway, reporting an increase in filed cases of child sexual abuse to the police during the pandemic compared to the years preceding the pandemic, especially in the capitol city (Oslo Police Derpartment, 2022).

That said, the 1.5 percentage point increase is relatively small, and even a small increase in absolute numbers can represent a significant change because the present sample size is large. However, from an epidemiological perspective even a 1.5 percentage point increase entail a considerable number of individuals in the age group included. It is also difficult to assess whether yearly fluctuations like presently observed for sexual abuse by an adult is normal despite a pandemic, and it is thus impossible to ascribe the increase to pandemic related factors. The same holds for the decrease in psychological violence. The observed decrease in prevalence during the first year of the pandemic, compared to the preceding year cannot entirely be seen as a consequence of the degree of preventive measures imposed. However, the slightly higher rate of psychological violence reported by adolescents living in more densely populated areas with more restrictions over a longer period of time, may indicate that mitigation measures have taken its toll in areas more affected by the COVID-19 pandemic in Norway. At the same time, we do not have data to investigate whether these differences in psychological violence in large urban versus smaller cities existed also before the pandemic.

The stability, and to some degree reduction in violence and abuse during the pandemic might be ascribed to at least two different factors. In Norway the rates of Covid infections and deaths were low, and there were significant periods of time with few measures to mitigate the spread of the virus.. In particular, parts of the country more distant to the metropolitan area of the capital city were almost non-affected by deaths, illness, and thus local restrictions. Although almost one third of Norway’s citizens live in or around the capital city of Oslo, large parts of the country did not experience restrictions other than quarantine and isolation when suspecting contagion or being infected by the virus, respectively. Although unemployment rates rose dramatically during the pandemic, and affected some groups more than others (e.g., young people, people with no higher education (Bratsberg, Markussen, Røed, Raaum, Vigtel, & Eielsen, 2020), social welfare measures were immediately imposed in order to compensate those who temporarily or permanently were laid off. These factors may have buffered family stress and thus violence towards adolescents at home, as we know from pre-pandemic research that stability and predictability is important for families in stressful life situations.

The stability or decrease in exposure to violence may also be ascribed to the age group included in the present study. Although schools were partly closed and after school activities in some parts of the country reduced, or periodically were put on halt, adolescents were allowed to meet and could leave the house unless they were ill and had to isolate. According to the United Nations Convention of the Rights of the child (Nations, 1989) and Norwegian legislation, the Norwegian government had a “the best interest of the child” policy when measures were implemented. An expert group of policy makers and public health experts was established under the Directorate for Child and Family Affairs in Norway, regularly monitoring the situation for children, adolescents and minorities in Norway. The Children’s ombudsman and other human rights institutions in Norway were proactive in constantly reminding politicians and policy makers of the importance of protecting children from the most invasive mitigation measures. In sum, these different factors might be associated with the relative stability in rates of violence when comparing the year before the pandemic to the first year of the pandemic.

### Strengths and limitations

The prospective longitudinal design with pre-pandemic baseline data, allowing for direct comparisons with the adolescents’ self-reported violence exposure one year prior to the Covid-19 pandemic, is a strength of this study. The study therefore makes an important addition to the body of literature that is scrutinizing the effects of the pandemic and the related restrictions on violence and abuse.

Due to the school-based survey design we may unintentionally have missed the most marginalized groups of adolescents, who we know from previous research may be more subject to violence. As such, we may have underestimated the actual rates of violence. That said, we would assume this bias would be present both before and during the pandemic, and as such not necessarily influencing the change or lack thereof. We were not able to include younger children in this survey, due to the study design and ethical permissions. Other research with baseline data from before the pandemic should consider the situation for younger children, as they are more reliant upon parents capable of taking care. Also, family and care related stress may be higher in families with smaller children, and where parents had to combine home office arrangements and care taking of younger children.

## Conclusion

The present study represents one of very few studies based on representative sample of adolescent self-report data on violence and sexual abuse exposure before and during the pandemic. The study findings indicate a slight increase in sexual abuse perpetrated by an adult, and a decrease in psychological violence by a caregiver at home during the pandemic compared to what was reported the year before the pandemic. Known risk factors for child abuse were also revealed, and the odds for abuse during the pandemic were somewhat higher for the identified risk groups compared to pre-pandemic odds ratios. Overall, the present study underscores the importance of monitoring child abuse rates in the adolescent population on a regular basis, as it is important for informing policy makers and service providers. It is also evident that prioritizing children and their needs during a pandemic is associated with no overall increase in abuse rates during the pandemic. At the same time, a focus on risk groups seems paramount as the risk for abuse increases for certain groups of adolescents during a pandemic.

## Data Availability

All data produced in the present study are available upon reasonable request to the authors and ethical approval by appropriate review board.

## Acknowledgements

We would like to thank the schools and students for taking time to answer this survey.

